# Predictors of Pregnancy-Related Anemia: A Logistic Regression Study at a Maternity Facility in Ghana

**DOI:** 10.64898/2026.07.16.26358280

**Authors:** Richard Yaw Kusi, Francis Anyan, Gerald Ohene Agyekum

**Affiliations:** Department of Mathematics, Tarleton State University, Stephenville, Texas, USA; Department of Statistical Sciences, Kumasi Technical University, Kumasi, Ghana

**Keywords:** anemia in pregnancy, logistic regression, gestational age, hemoglobin, predictive model

## Abstract

**Background:** Anemia during pregnancy remains a major public health concern, particularly in low- and middle-income countries, where it contributes substantially to maternal and neonatal morbidity and mortality. Identifying women at increased risk is essential for timely intervention and improved pregnancy outcomes.

**Objective:** This study aimed to identify significant predictors of anemia among pregnant women using logistic regression and to evaluate the association between selected clinical characteristics and anemia status.

**Methods:** A retrospective study was conducted using secondary data obtained from Ayiwa Memorial Maternity Home in the Suame Municipality of the Kumasi Metropolis, Ashanti Region, Ghana, covering the period 2018 to 2023. A sample of 396 pregnant women with complete information on hemoglobin status and candidate predictor variables was analyzed. The outcome variable was whether a woman had ever had anemia (hemoglobin concentration below 11 g/dL). Candidate predictors included maternal age, diastolic and systolic blood pressure, parity, height, weight, gestational age, sickle cell status, employment, and education. Multivariable logistic regression was fitted, with multicollinearity among predictors assessed using variance inflation factors (VIF) and tolerance statistics. Model performance was evaluated using the Hosmer-Lemeshow test, Nagelkerke *R*^2^, and the area under the receiver operating characteristic curve (AUC).

**Results:** Of the 396 pregnant women studied, 69.44% were classified as anemic. In the multivariable logistic regression model, gestational age, maternal weight, and diastolic blood pressure were significantly associated with anemia status (*p <* 0.05). Higher gestational age was associated with increased odds of anemia (AOR = 1.124, 95% CI: 1.073–1.180), while higher weight (AOR = 0.975, 95% CI: 0.955–0.995) and higher diastolic blood pressure (AOR = 0.964, 95% CI: 0.934–0.993) were associated with reduced odds of anemia. Systolic blood pressure, height, and education were not significantly associated with anemia in the final model. The model showed acceptable discriminatory ability (AUC = 0.752, 95% CI: 0.694–0.810) and adequate calibration (Hosmer-Lemeshow *χ*^2^ = 13.323, *p* = 0.101), explaining approximately 23.9% of the variation in anemia status (Nagelkerke *R*^2^ = 0.239), with high specificity (91.8%) but limited sensitivity (36.5%).

**Conclusion:** Gestational age, maternal weight, and diastolic blood pressure were identified as significant predictors of anemia during pregnancy in this population. These findings support intensifying anemia screening as pregnancy advances and incorporating nutritional assessment into routine antenatal care. However, given the model’s limited sensitivity, clinical risk models cannot replace routine laboratory hemoglobin testing. Further prospective, multi-center studies incorporating dietary, infectious, and socioeconomic determinants are recommended to validate these findings.

## 1 Introduction

The World Health Organization (WHO) defines anemia as having a hemoglobin concentration of less than 11 grams per deciliter (g/dL). In 2019, the incidence of anemia among women of reproductive age was 29.9% worldwide, affecting about 500 million women aged 15 to 49 (Petraglia et al., 2024). Among pregnant women, it was 36.5%, while among non-pregnant women, it was 29.6%. Globally, the prevalence has remained steady despite a slight decrease since 2000 (Petraglia et al., 2024).

Anemia is the most common nutritional problem disproportionately affecting women of reproductive age in all settings due to women’s physiologic state of menstruation, pregnancy, and lactation (Mare et al., 2023; Raut et al., 2022). Although the risk of exposure to anemia differs across preconception, pregnancy, and lactation periods, its occurrence throughout women’s reproductive life is associated with poor maternal, newborn, and child health outcomes (Mare et al., 2023; Shah et al., 2022; Uzunov et al., 2022; Liu et al., 2022). In 2019, anemia affected half a billion (30%) women of reproductive age and 37% of pregnant women globally, with Africa ranked second for anemia (62.3%) and severe anemia (1.8%) (Tirore et al., 2024).

The risk of inactivity, maternal sickness, and death is increased by severe anemia. The general health of pregnant women becomes endangered by anemia because it leads to fatigue and reduced physical and cognitive abilities and decreased functional capacity. Iron deficiency is the leading cause of anemia, accounting for an estimated 50% of cases in women globally, despite the disease’s complex character (Tirore et al., 2024). Gestational anemia results in an increased risk of antepartum, intrapartum, and postpartum maternal hemorrhage and subsequent blood transfusions, pregnancy-induced hypertension, prolonged and induced labor, cesarean birth, puerperal infections, maternal shock, longer hospitalization and critical care admission, and mortality (Mare et al., 2023; Shi et al., 2022). Likewise, the risk of poor neonatal outcomes such as low birth weight, small-for-gestational-age, preterm birth, low Apgar score, stillbirth, and neonatal and perinatal mortality is higher among neonates and children born to anemic mothers (Mare et al., 2023; Shah et al., 2022; Uzunov et al., 2022; Shi et al., 2022).

In Sub-Saharan Africa, the prevalence of anemia is 43%, with the highest prevalence in the Western African region (51%), and anemia was found to be a severe public health problem (prevalence ≥ 40%) in 18 countries (Mare et al., 2023; Stevens et al., 2022; Kinyoki et al., 2021; Correa-Agudelo et al., 2021). Different interventions like iron-folic acid supplementation and food fortification have reduced anemia, but progress remains slow. Research needs to increase worldwide to better understand the risk factors for anemia in pregnant women to achieve better health outcomes. This research aims to identify potential risk factors for anemia among pregnant women in Suame Municipality, Ghana.

## 2 Literature Review

Gebremedhin et al. (2014) investigated the prevalence and contributing factors of anemia in pregnant women in rural Sidama, Southern Ethiopia, showing that low socioeconomic position and insufficient dietary iron intake were important risk factors. Armah-Ansah (2023) examined factors contributing to anemia in Mali’s reproductive-age population, identifying education, obesity, and wealth as significant correlates.

In a large retrospective cohort study in China involving over 18 million pregnant females, Shi et al. (2022) found that 17.78% were diagnosed with anemia during pregnancy and demonstrated that severe anemia was associated with placental abruption, preterm birth, and postpartum hemorrhage. Multi-country analyses of Demographic and Health Survey (DHS) data across Sub-Saharan Africa by Mare et al. (2023) and Tirore et al. (2024) revealed that illiteracy, household poverty, non-attendance of antenatal care, and underweight status were positively linked with higher anemia odds.

In Cambodia, Um et al. (2023) showed that pregnant women in their second and third trimesters were significantly more likely to have anemia compared to those in their first trimester. In Ethiopia, Animut and Berhanu (2022) found that antenatal care visits and iron supplementation were key protective factors. Finally, Nadhifa et al. (2023) documented a relationship between gestational weight gain and anemia status, emphasizing the role of maternal nutritional reserves.

## 3 Data and Methods

### 3.1 Data Source and Sample

The data for this study were obtained from Ayiwa Memorial Maternity Home, Suame, within the Kumasi Metropolis in Ghana, covering clinical records from 2018 to 2023. A retrospective analytic sample of 396 pregnant women with complete medical records was compiled.

The outcome variable was binary: whether a pregnant woman had ever been diagnosed with anemia during pregnancy (hemoglobin *<* 11 g/dL, coded as 1) or not (coded as 0). The initial candidate predictor variables evaluated were maternal age, diastolic blood pressure, systolic blood pressure, parity, height, weight, gestational age, sickle cell status, employment status, and education level.

### 3.2 Statistical Analysis

Statistical analyses were performed using R software (version 4.5.1; R Core Team, 2025). Multivariable logistic regression was used to evaluate predictors of anemia. The standard logit model formulation is specified as:

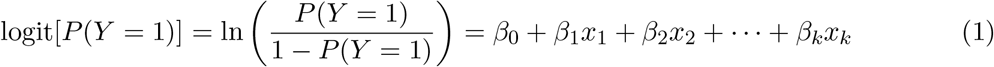

where *P* (*Y* = 1) is the probability of having anemia, *x*_1_, …, *x*_*k*_ are the predictor variables, *β*_0_ is the intercept, and *β*_1_, …, *β*_*k*_ are the estimated log-odds regression coefficients. Regression coefficients were exponentiated to yield Adjusted Odds Ratios (AOR = *e*^*β*^) with 95% confidence intervals.

Multicollinearity among continuous predictors was evaluated using Pearson correlation matrix, Variance Inflation Factors (VIF), and tolerance statistics, consistent with diagnostic procedures used in logistic regression modeling for clinical risk prediction (Kusi, 2026). Categorical predictors (e.g., education) were dummy coded before diagnostic checking and model entry. Model goodnessof-fit was assessed using the Hosmer-Lemeshow test and Nagelkerke *R*^2^. Model discrimination was evaluated using Receiver Operating Characteristic (ROC) curve analysis and Area Under the Curve (AUC), alongside sensitivity, specificity, and classification accuracy.

## 4 Results

Among the 396 pregnant women in the study sample, 275 (69.44%) were classified as anemic based on a hemoglobin concentration below 11 g/dL.

Table 1 displays the clinical characteristics of participants. The mean age was 29.83 *±* 5.78 years. Mean systolic and diastolic blood pressures were 109.59 mmHg and 68.09 mmHg, respectively. Average gestational age at presentation was 15.93 weeks (range: 4 to 40 weeks).

**Table 1:** Descriptive Statistics of the Continuous Variables (*N* = 396).

| Variables | Mean | Standard Deviation | Minimum | Maximum |
| --- | --- | --- | --- | --- |
| Age (years) | 29.83 | 5.78 | 14 | 57 |
| Diastolic BP (mmHg) | 68.09 | 11.57 | 40 | 130 |
| Systolic BP (mmHg) | 109.59 | 13.97 | 80 | 200 |
| Parity | 1.67 | 1.51 | 0 | 8 |
| Height (cm) | 160.71 | 7.41 | 137 | 192 |
| Weight (kg) | 69.06 | 14.20 | 43 | 115 |
| Gestational Age (weeks) | 15.93 | 11.34 | 4 | 40 |

**Table 2:** Correlation Matrix and Multicollinearity Diagnostics.

| Predictor | Diastolic BP | Systolic BP | Height | Weight | Gestation | VIF | Tolerance |
| --- | --- | --- | --- | --- | --- | --- | --- |
| Diastolic BP | 1.000 | 0.630 | 0.063 | 0.230 | -0.068 | 1.271 | 0.786 |
| Systolic BP | 0.630 | 1.000 | 0.089 | 0.204 | -0.006 | 1.254 | 0.798 |
| Height | 0.063 | 0.089 | 1.000 | 0.165 | 0.023 | 1.027 | 0.973 |
| Weight | 0.230 | 0.204 | 0.165 | 1.000 | 0.127 | 1.091 | 0.916 |
| Gestation | -0.068 | -0.006 | 0.023 | 0.127 | 1.000 | 1.062 | 0.941 |
| Education | — | — | — | — | — | 1.022 | 0.978 |

*Note:* Education was dummy-coded before model evaluation; correlation values with continuous parameters are excluded from the Pearson matrix.

**Table 3:** Multivariable Logistic Regression Analysis.

| Variables | Adjusted Odds Ratio | 95% Confidence Interval |  | *P*-value |
| --- | --- | --- | --- | --- |
|  |  | Lower | Upper |  |
| Diastolic BP | 0.964 | 0.934 | 0.993 | 0.018 |
| Systolic BP | 1.019 | 0.994 | 1.044 | 0.136 |
| Height | 1.017 | 0.981 | 1.053 | 0.362 |
| Weight | 0.975 | 0.955 | 0.995 | 0.019 |
| Gestation | 1.124 | 1.073 | 1.180 | 0.001 |
| Education | 0.480 | 0.177 | 1.225 | 0.134 |

*Note:* Initial candidate variables age, parity, sickle cell status, and employment were excluded during variable selection ( $p > 0.10$ ).

Multicollinearity among predictors was assessed using variance inflation factors (VIF) and tolerance values. VIF quantifies how much the variance of regression coefficient estimates increases due to correlations among predictors (Agyekum et al., 2023). Correlations among continuous predictors ranged from weak to moderate (*r* = −0.068 to 0.630). All predictors exhibited VIF values *<* 5 and tolerance values *>* 0.20, confirming that multicollinearity was not present.

In the adjusted model, gestational age, maternal weight, and diastolic blood pressure were statistically significant predictors (*p <* 0.05). Each additional week of gestational age increased anemia odds by 12.4% (AOR = 1.124, 95% CI: 1.073–1.180). Higher weight (AOR = 0.975, 95% CI: 0.955–0.995) and higher diastolic BP (AOR = 0.964, 95% CI: 0.934–0.993) were associated with significantly reduced odds of anemia.

The model demonstrated acceptable overall discrimination (AUC = 0.752) and adequate calibration (Hosmer-Lemeshow *p* = 0.101). However, while specificity was high (91.8%), sensitivity was low (36.5%), indicating that the model missed a notable proportion of true anemia cases.

**Table 4:**
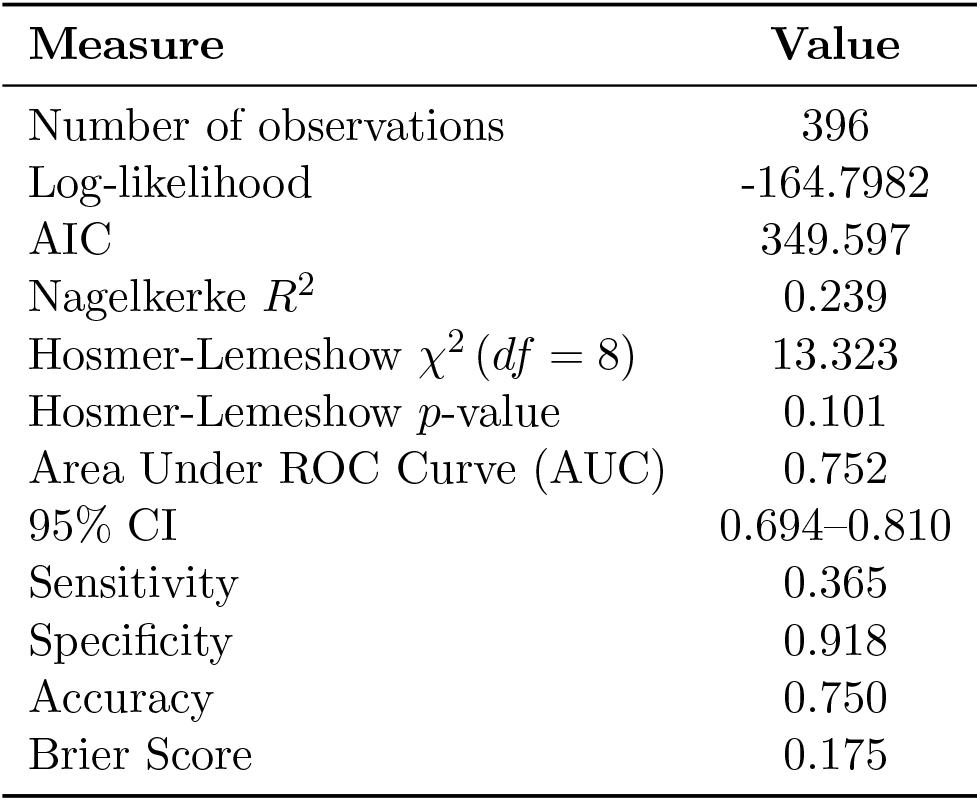
Model Fit and Predictive Performance.

## 5 Discussion

### 5.1 Key Findings and Physiological Content

Among the 396 pregnant women evaluated, the prevalence of anemia was 69.44%, substantially exceeding global estimates (36.5%) and underscoring a severe localized public health burden. Gestational age showed a positive association with anemia risk (AOR = 1.124). This aligns with progressive plasma volume expansion relative to red blood cell mass production as gestation advances, causing a physiological dilutional effect.

Maternal weight was inversely associated with anemia risk (AOR = 0.975). Higher weight may reflect greater nutritional reserves, whereas underweight status frequently correlates with micronutrient deficiencies.

The inverse relationship observed for diastolic BP (AOR = 0.964) requires cautious interpretation. Women experiencing larger plasma volume expansion often exhibit lower BP alongside a greater dilutional drop in hemoglobin. This finding should be treated as hypothesis-generating rather than an indicator that elevated BP is protective.

### 5.2 Non-Significant Predictors and Study Limitations

Systolic blood pressure, height, and education were not statistically significant in the final model. The lack of significant association for education contrasts with population-wide DHS surveys in Sub-Saharan Africa. This divergence likely stems from sample homogeneity within a single urban facility.

#### Study Limitations

1. **Single-Center Design:** Secondary clinical records from a single facility limit generalizability across Ghana.
2. **Unmeasured Confounders:** Crucial factors such as dietary diversity, iron/folate compliance, malaria status, intestinal parasites, and detailed income were unavailable.
3. **Low Model Sensitivity:** A sensitivity of 36.5% indicates that clinical screening algorithms based solely on routine physical measurements cannot substitute for standard laboratory hemoglobin assays.

## 6 Conclusion

Gestational age, maternal weight, and diastolic blood pressure were identified as significant predictors of anemia during pregnancy in this Ghanaian cohort. While these clinical metrics provide valuable contextual risk indicators, the model’s low sensitivity emphasizes that clinical models must complement, rather than replace, routine blood hemoglobin testing. Antenatal care programs should prioritize nutritional monitoring and regular laboratory hemoglobin screening throughout gestation.

## Declarations

### Data Availability

Data are available upon reasonable request from the corresponding author.

### Ethics Approval

Ethical approval was granted by the Kumasi Technical University Research Ethics Committee.

